# Usefulness of the Short Form-8 (SF-8) for chronic pain in the orofacial region

**DOI:** 10.1101/2023.05.15.23289230

**Authors:** Aiji Sato (Boku), Tatsuya Tokura, Hiroyuki Kimura, Mikiko Ito, Shinichi Kishi, Takashi Tonoike, Norio Ozaki, Yumi Nakano, Saori Nakano, Hiroshi Hoshijima, Masahiro Okuda

**Affiliations:** Department of Anesthesiology, School of Dentistry, Aichi Gakuin University, 2-11 Suemori-dori, Chikusa-ku, Nagoya, 464-8651, Japan; Department of Psychiatry, Nagoya University Graduate School of Medicine, 65 Tsurumai-cho, Showa-ku, Nagoya, 466-8650, Japan; Department of Oral and Maxillofacial Surgery, School of Dentistry, Aichi Gakuin University, 2-11 Suemori-dori, Chikusa-ku, Nagoya, 464-8651, Japan; Faculty of Psychological and Physical Sciences, Health Service Center, Aichi Gakuin University, 12 Araike Iwasaki-cho, Nisshin, 470-0195, Japan; Institute for Glyco-core Reserch (iGcORE), Nagoya University Graduate School of Medicine, 65 Tsurumai-cho, Showa-ku, Nagoya, 466-8650, Japan; Department of Psychology and Human Relations, Nanan University, 18 Yamazaki-cho, Showa-ku, Nagoya, 466-8673, Japan; Aichi PFS Association, 1-21-35 Osu, Naka-ku, Nagoya, 460-0011, Japan; Division of Dento-oral Anesthesiology, Tohoku University Graduate School of Dentistry, Seiryomachi 4-1, Aoba-ku, Sendai, 980-0000, Japan

## Abstract

Given that chronic pain has become a major problem in recent years, affecting approximately 30% of the general population, this study used the Short Form-8 (SF-8) Japanese version to investigate (1) the quality of life (QOL) of patients with burning mouth syndrome (BMS) or persistent idiopathic facial pain (PIFP) (compared to a Japanese control group) and (2) whether the therapeutic intervention improves the QOL and reduced pain (comparison between 0 and 12 weeks) of patients with BMS or PIFP. A total of 63 patients diagnosed with either BMS (n = 45) or PIFP (n = 18) were included in this study. The diagnostic criteria for BMS and PIFP were established based on the 3rd edition of the International Classification of Headache Disorders. Our study results showed that while Physical Component Summary (PCS) in patients with BMS or PIFP improved with treatment, it did not improve to the national standard value (NSV) after 12 weeks of intervention. In contrast, Mental Component Summary (MCS) improved to the same level as NSV after 12 weeks of intervention. Therefore, we found that therapeutic intervention improves MCS and reduces pain; however, improving PCS takes time.

## Introduction

Chronic pain has become a major problem in recent years, affecting approximately 30% of the general population [1]. The majority of these patients are difficult to treat, and even when they can be treated, 50% of them experience only partial improvement and reduced quality of life (QOL) [2,3]. However, understanding chronic pain is difficult; thus, it is underdiagnosed and under-treated because pain cannot be commensurate with the organic abnormality [4,5]. Chronic pain can be a comorbidity of mental illness such as depression and can also affect various aspects of patients’ daily lifestyles, such as housework and employment, which immensely lowers their quality of life [6].

Chronic pain in the orofacial region includes various conditions, such as burning mouth syndrome (BMS), persistent idiopathic facial pain (PIFP), and nonorganic temporomandibular joint disorder. In particular, BMS and PIFP are commonly encountered in daily clinical practice [7]. Since these patients often complain of physical symptoms only, which are the main complaints, establishing an accurate diagnosis, identifying the treatment, and evaluating the degree of improvement are difficult even after the treatment intervention.

The concept of health-related quality of life (HRQOL) has been generally used as a multidimensional assessment of how disease and treatment affect a patient’s sense of overall function and well-being [8]. It is also an inclusive concept based on the patient’s subjective judgment. In other words, HRQOL quantifies the impact of an illness on performing activities of daily living (ADL). The Short Form-8 (SF-8) is a comprehensive, versatile, and practical tool globally used tool for measuring HRQOL [9], allowing comparison of normative values from large national surveys with results from more focused outcome studies. The SF-8 is based on the SF-36, a 36-item version of the rating scale, but is more convenient because it provides results equivalent to the SF-36. Although the authors provided treatment interventions for many patients with chronic pain in the orofacial region, pre- and post-treatment evaluations were based on conventional clinical diagnostic evaluations based on physicians and dentists and were inadequate from the patient’s viewpoint. In addition to pain reduction, the goal of treatment is to improve the QOL; however, to the best of the authors’ knowledge, no previous reports evaluated the QOL at pre- and post-treatment intervention for chronic pain in the orofacial region.

In this study, the SF-8 Japanese version [10] was used to investigate 1) the QOL of patients with BMS or PIFP (compared to a Japanese control group) and 2) whether the therapeutic intervention improves the QOL and reduced pain (comparison between 0 and 12 weeks).

## Materials and methods

### Ethical guidelines

This study was conducted per the principles laid out in the Declaration of Helsinki and was approved by the ethical review committee of Nagoya University Graduate School of Medicine (no. 234, 234-2, 2004-0234-2 and 2004-0234-3) and the ethical committee of the School of Dentistry, AichiLJGakuin University (no. 41). Every effort was made to protect the patients’ confidentiality and personal information. All participants provided written informed consent.

### Study design and patients

The study was initiated on May 10, 2010 and patients were recruited between May 10, 2010 and April 21, 2021. A total of 63 patients diagnosed with either BMS (n = 45) or PIFP (n = 18) who were treated at the Liaison Clinic, Department of Oral and Maxillofacial Surgery, Aichi Gakuin University Dental Hospital (coordinated with the Department of Psychiatry, Nagoya University Graduate School of Medicine) and who consented to participate were included in this study. The diagnostic criteria for BMS and PIFP were established based on the 3rd edition of the International Classification of Headache Disorders (Table 1) [11].

**Table 1.**
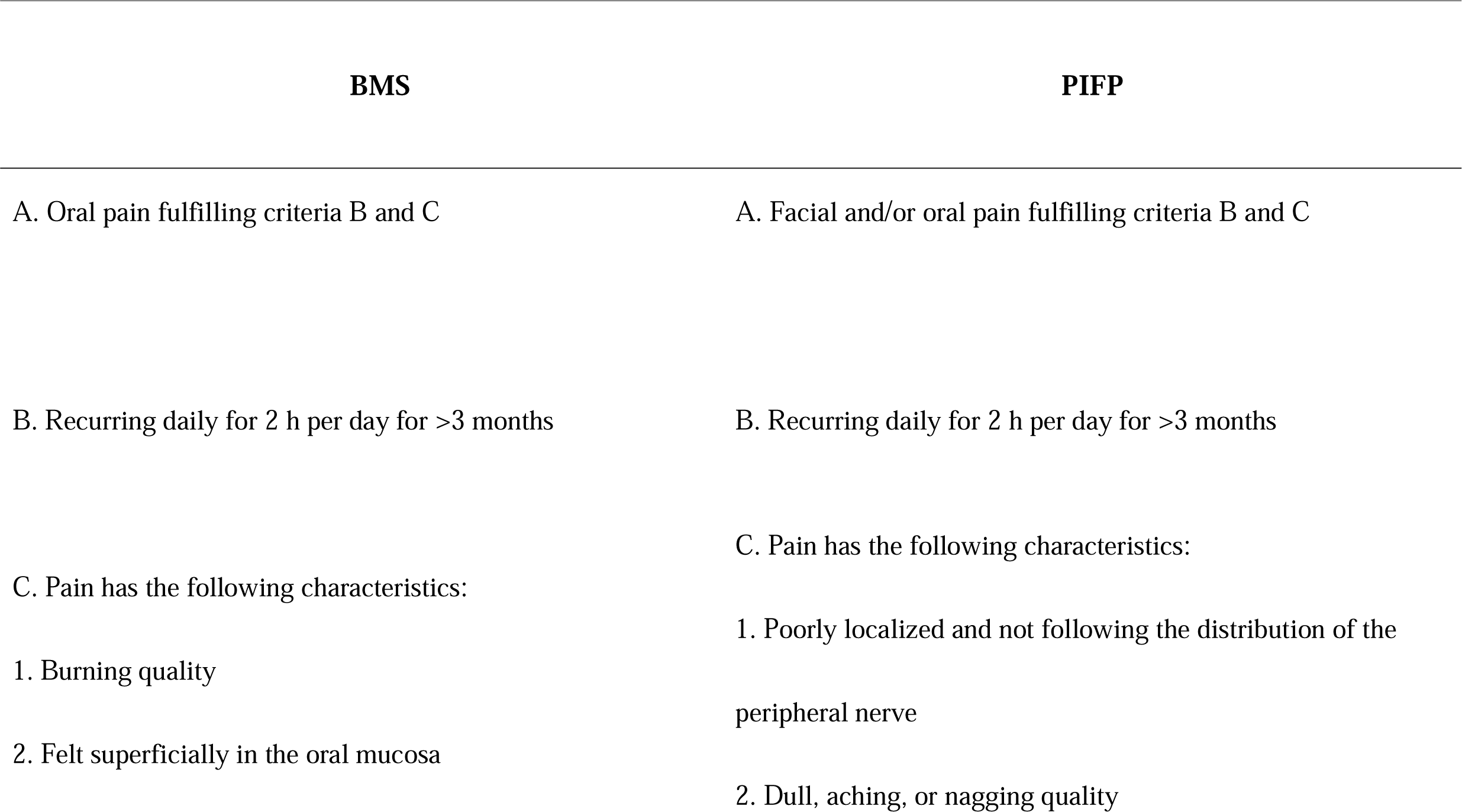
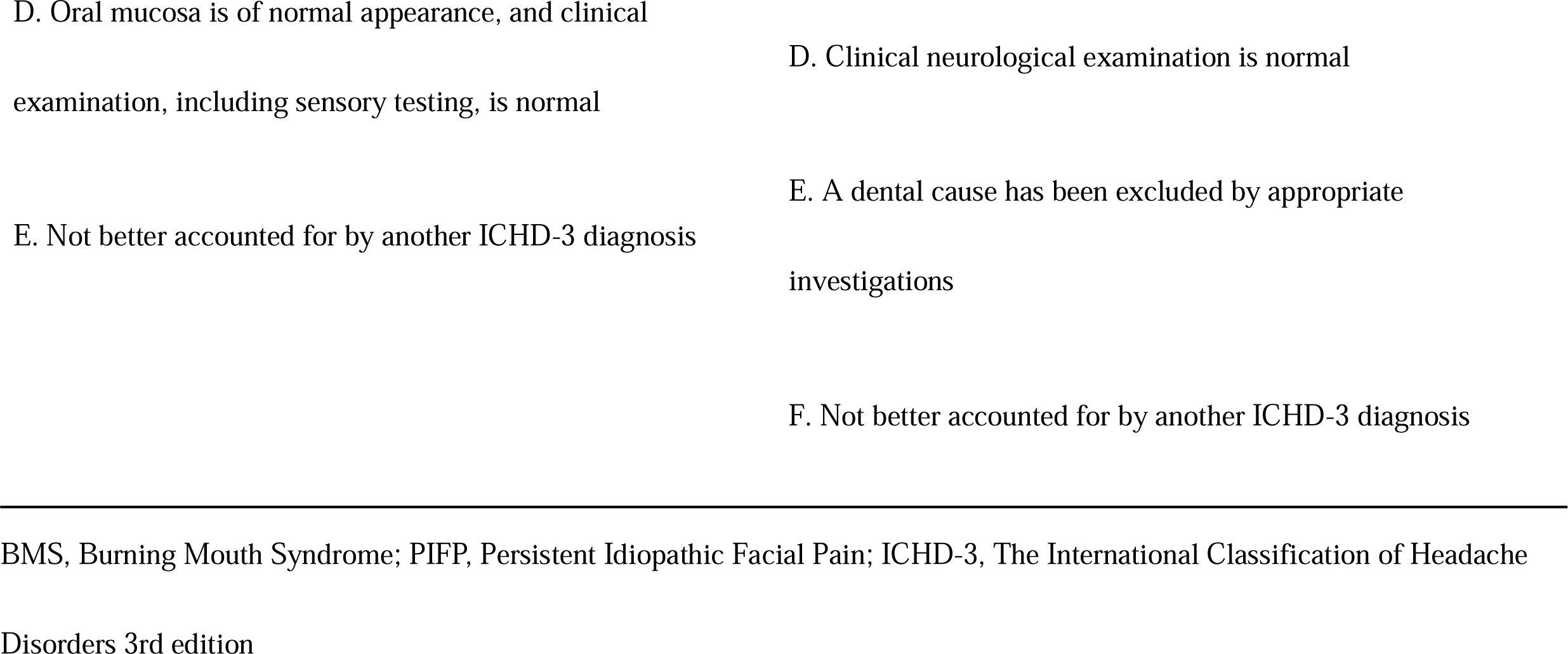
Diagnostic Criteria of BMS and PIFP Based on the 3rd Edition of the International Classification of Headache Disorder.

A trained psychiatrist performed psychiatric evaluations on all patients using the Diagnostic and Statistical Manual of Mental Disorders, Fifth Edition (DSM-5) [12]. Patients who visited our liaison clinic before 2013 were diagnosed using the Diagnostic and Statistical Manual of Mental Disorders, Fourth Edition, Text Revision (DSM-IV-TR) [13]. These patients’ psychiatric diagnoses were recategorized based on DSM-5 by confirming each patient’s clinical record.

To make a psychiatric diagnosis based on DSM-5, a structured clinical interview was conducted. The psychiatrist and dentist are providing treatment in the same clinic, and the psychiatric and dental diagnoses were performed in the same place and at the same time. Regarding therapeutic intervention, serotonin noradrenaline reuptake inhibitors (SNRIs) are recently more commonly used than tricyclic antidepressants because of their fewer adverse effects although antidepressants have been widely used for the treatment of chronic pain. Our previous study reported that SNRI duloxetine is effective for chronic pain in the orofacial region [14-17]. Based on the above, duloxetine was also used in the present study and administered to patients with the above diagnosis from the initial visit (0 weeks). The initial dose of duloxetine is 20 mg once daily. At ≥2 weeks after initiating the administration, the dose was increased to a maximum of 40 mg once daily observing symptom changes and adverse effect expressions.

The collected data were anonymized, and the authors did not have access to information that could identify individual participants once it was stored.

### Outcome measures

As the main outcome, for patients who received the therapeutic intervention, the SF-8 at the initial visit and after 12 weeks of intervention were evaluated and compared with those of the Japanese control participants. The SF-8 consists of the following items: general health, physical function, daily role function (physical), physical pain, vitality, social function, mental health, and daily role function (mental) (Table 2) [9]. These eight items were scored using norm-based scoring (NBS: scoring based on the national standard value [NSV] [50]) for each item. Based on the eight items, two summary scores, “Physical Component Summary (PCS)” and “Mental Component Summary (MCS),” were calculated to indicate physical and mental health, respectively. For both PCS and MCS, higher scores indicate a higher quality of life. A group of healthy participants matched in the number with the patient group was also created from the NBS data and used as the NSV.

**Table 2.**
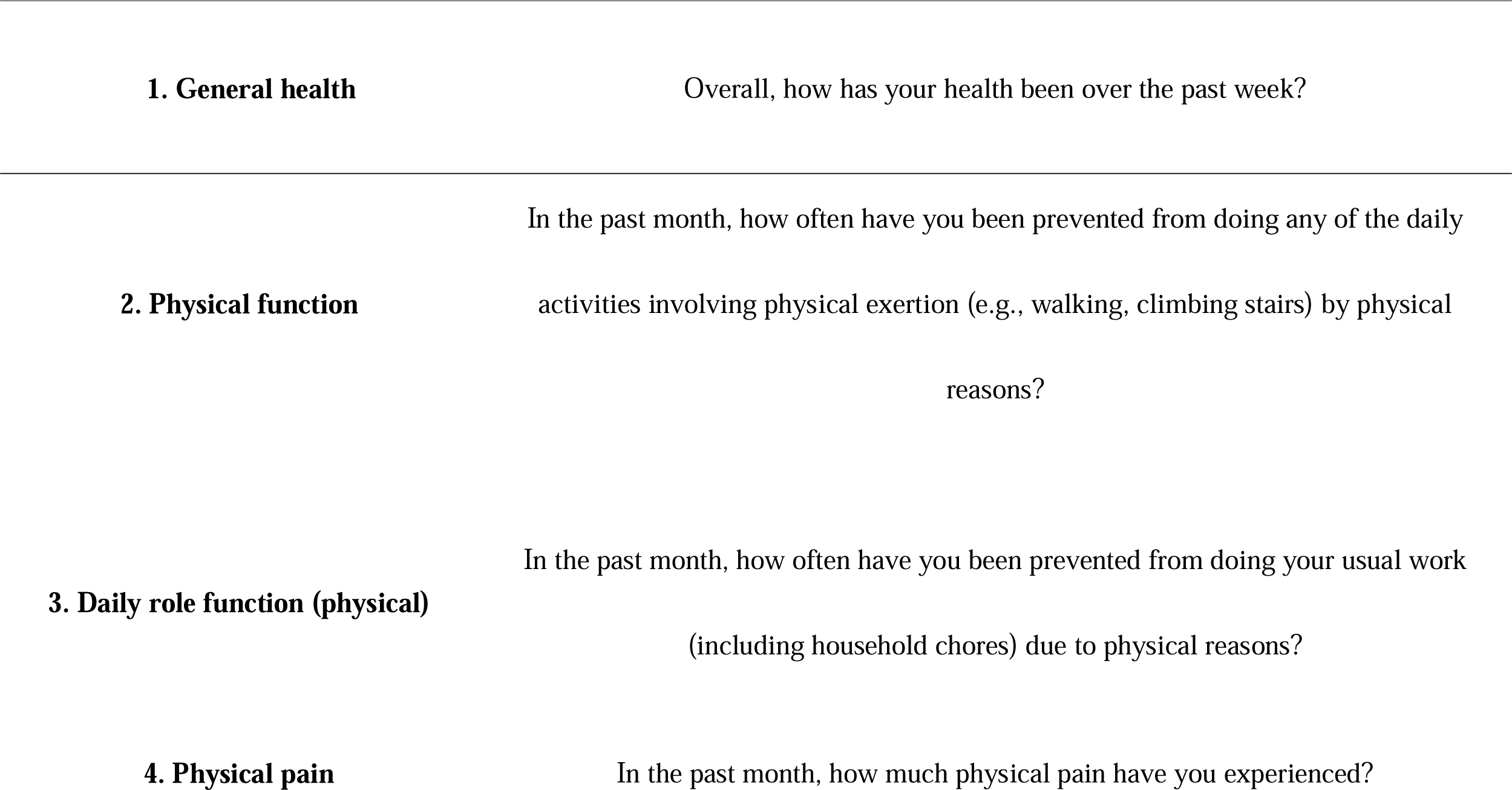
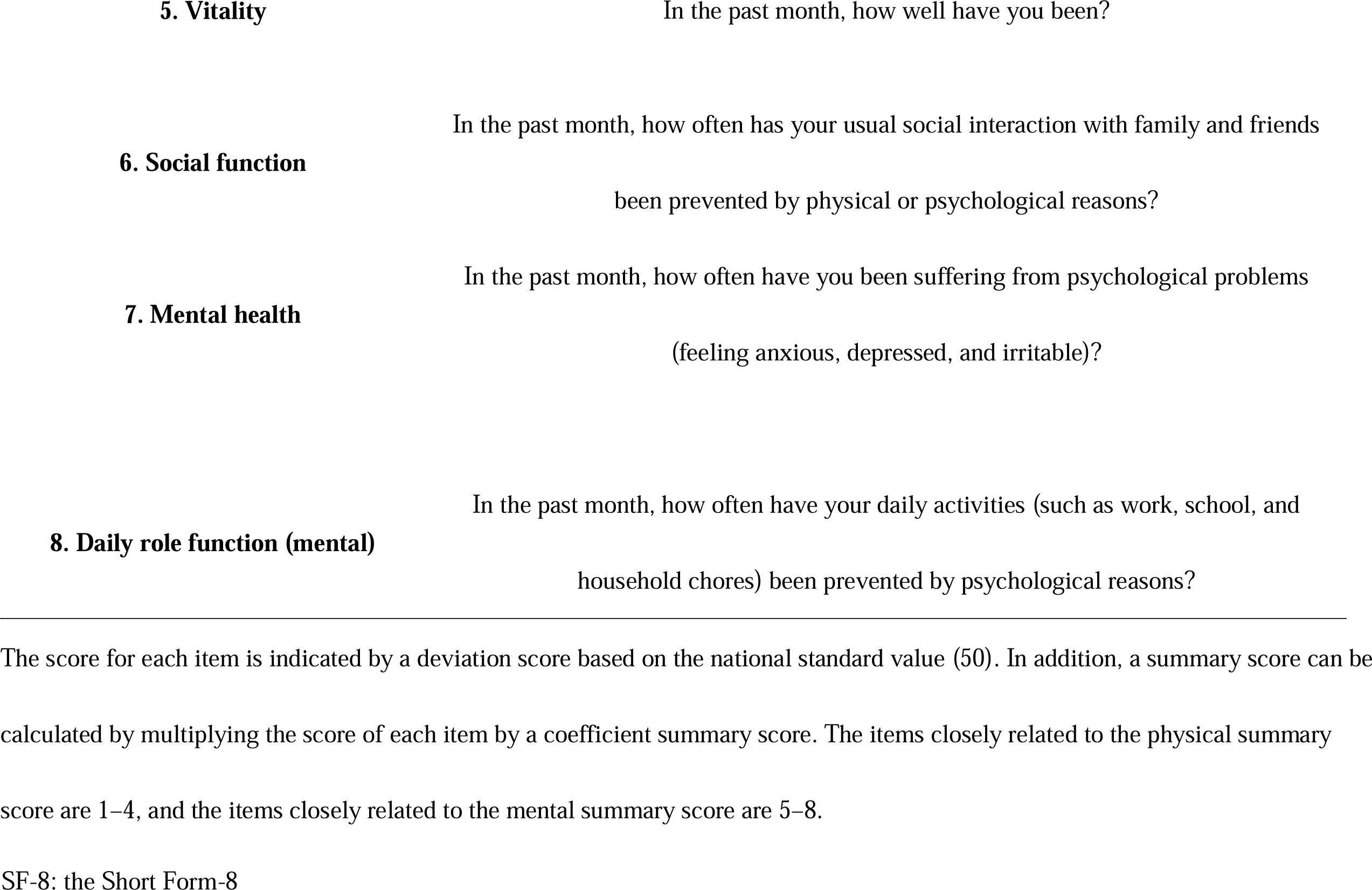
Component Concepts and Questions of SF-8.

Additionally, pain intensity was evaluated using a visual analog scale (VAS) value. To evaluate depression in 63 patients with BMS and PIFP, Beck’s depression inventory (BDI: cutoff value, ≤10) was used as a subjective index and the Hamilton depression rating scale (HDRS: cutoff value, ≤7), which uses semi-structured interviews by a trained psychiatrist, as a highly precise objective index [18,19].

### Statistical analysis

Data are expressed as median (interquartile range [IQR]) or number. Since the sample size is small and the data do not follow a normal distribution, we adopted the median value instead of the average value. For statistical testing, the Mann–Whitney U-test was used to compare differences between two independent groups for continuous variables. The two-sided statistical significance level was set at p ≤ 0.05. Statistical analysis of the recorded data was performed using IBM® SPSS® Statistics Ver26.

## Results

A total of 63 patients participated in the study. Table 3 depicts the demographic characteristics and psychiatric diagnosis.

**Table 3.**
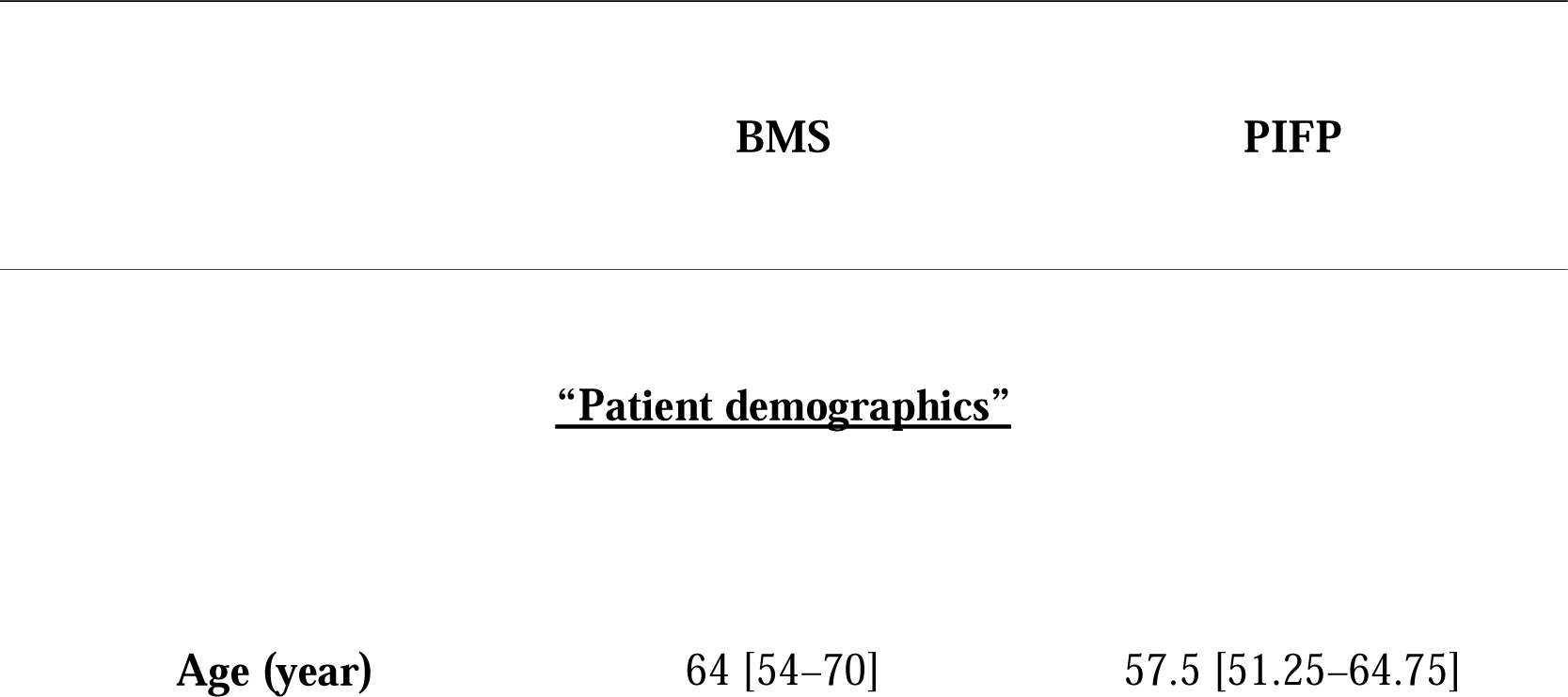
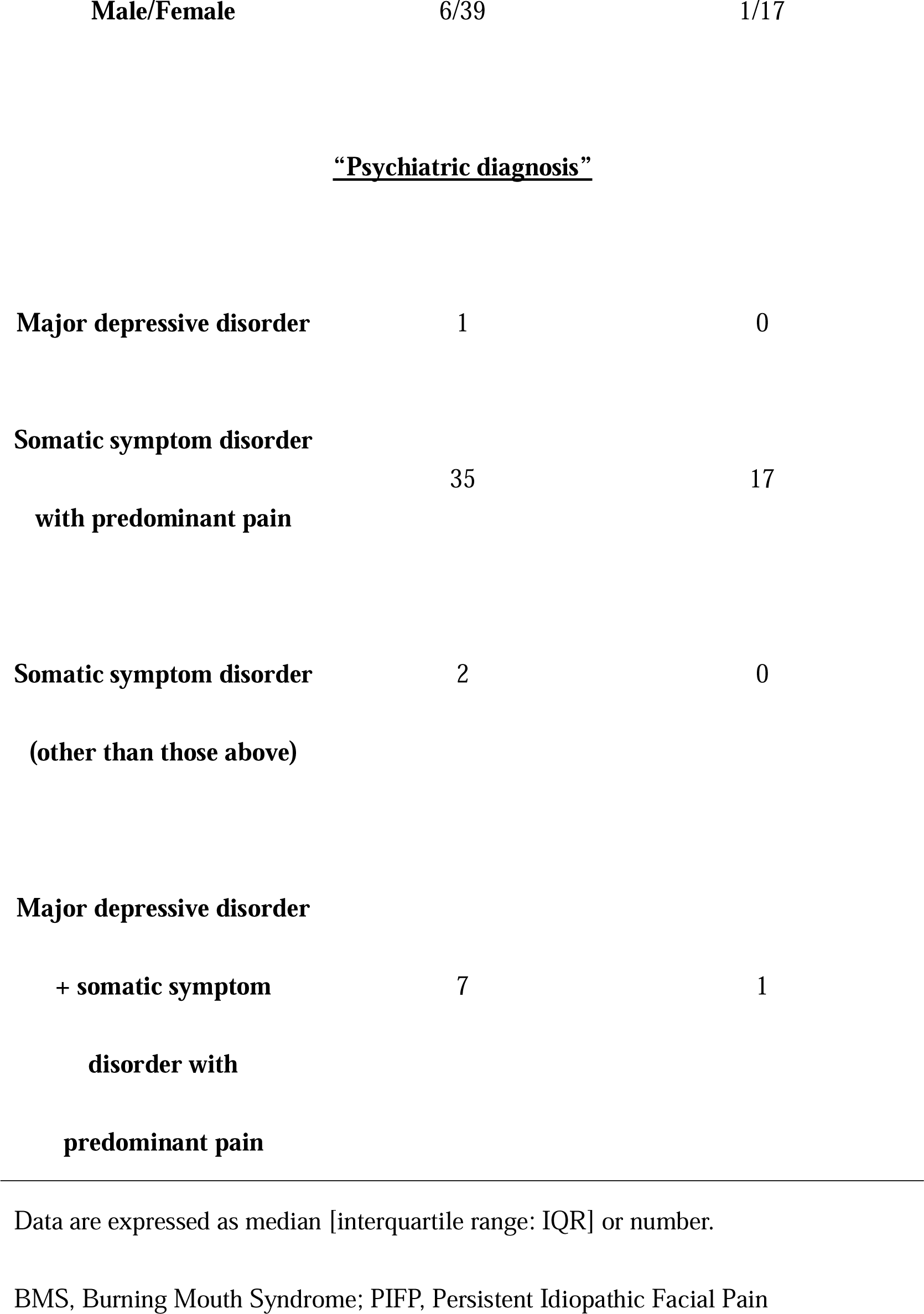
Patient Characteristics in this Study.

As shown in Fig 1, the SF-8 PCS was 41.3 [37.1–45.9] at 0 week, 45.3 [40.2–50.1] at 12 weeks, and the NSV was 50.28 [45.86–53.28]. Statistically significant differences were observed between 0 and 12 weeks, 0 week and NSV, and 12 weeks and NSV. The SF-8 MCS was 45.4 [38.4–49.6] at 0 week and 48.1 [44.4–52.3] at 12 weeks, and the NSV was 49.86 [45.96–53.49]. Statistically significant differences were observed between 0 week and 12 weeks and between 0 week and NSV. However, no statistically significant difference was observed between 12 weeks and NSV.

**Fig 1.**
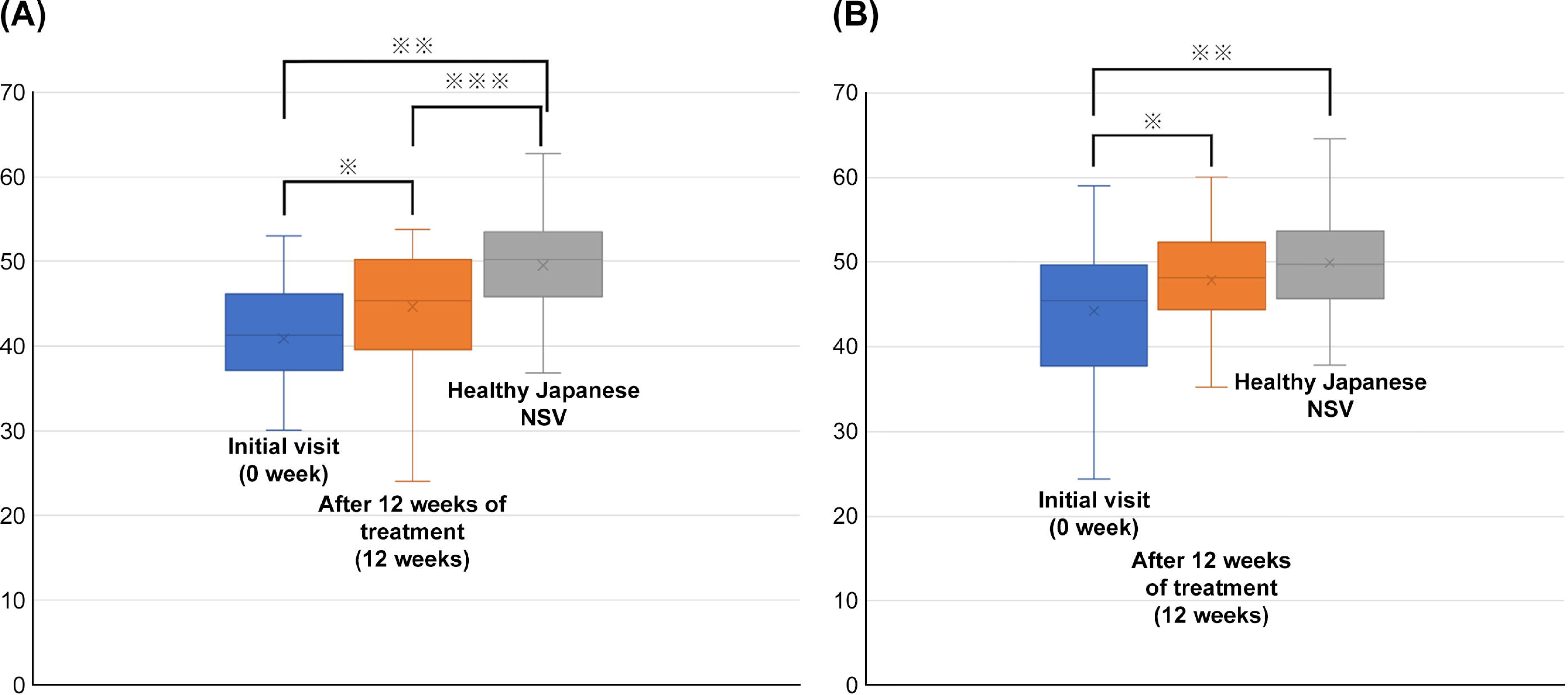
**Figure 1A. Comparison of SF-8 PCS between at initial visit, after 12 weeks of treatment, and healthy Japanese data.** ※P<0.01 (0week vs 12weeks), ※※ P<0.01 (0week vs NSV), ※※※P<0.01 (12weeks vs NSV) **Figure 1B. Comparison of SF-8 MCS between at initial visit, after 12 weeks of treatment, and healthy Japanese data.** ※P=0.01 (0week vs 12weeks) ※※P<0.01 (0week vs NSV)

Fig 2 shows the VAS, BDI, and HDRS between the initial visit and after 12 weeks of treatment. The VAS was 53 [32.5–77] at 0 week and 26 [8.5–46] at 12 weeks. The BDI was 12 [6.5–18.5] at 0 week and 6 [1.5–10.5] at 12 weeks. The HDRS was 6 [3–12] at 0 week and 2 [1–3] at 12 weeks. All parameters showed statistically significant differences between 0 and 12 weeks.

**Fig 2.**
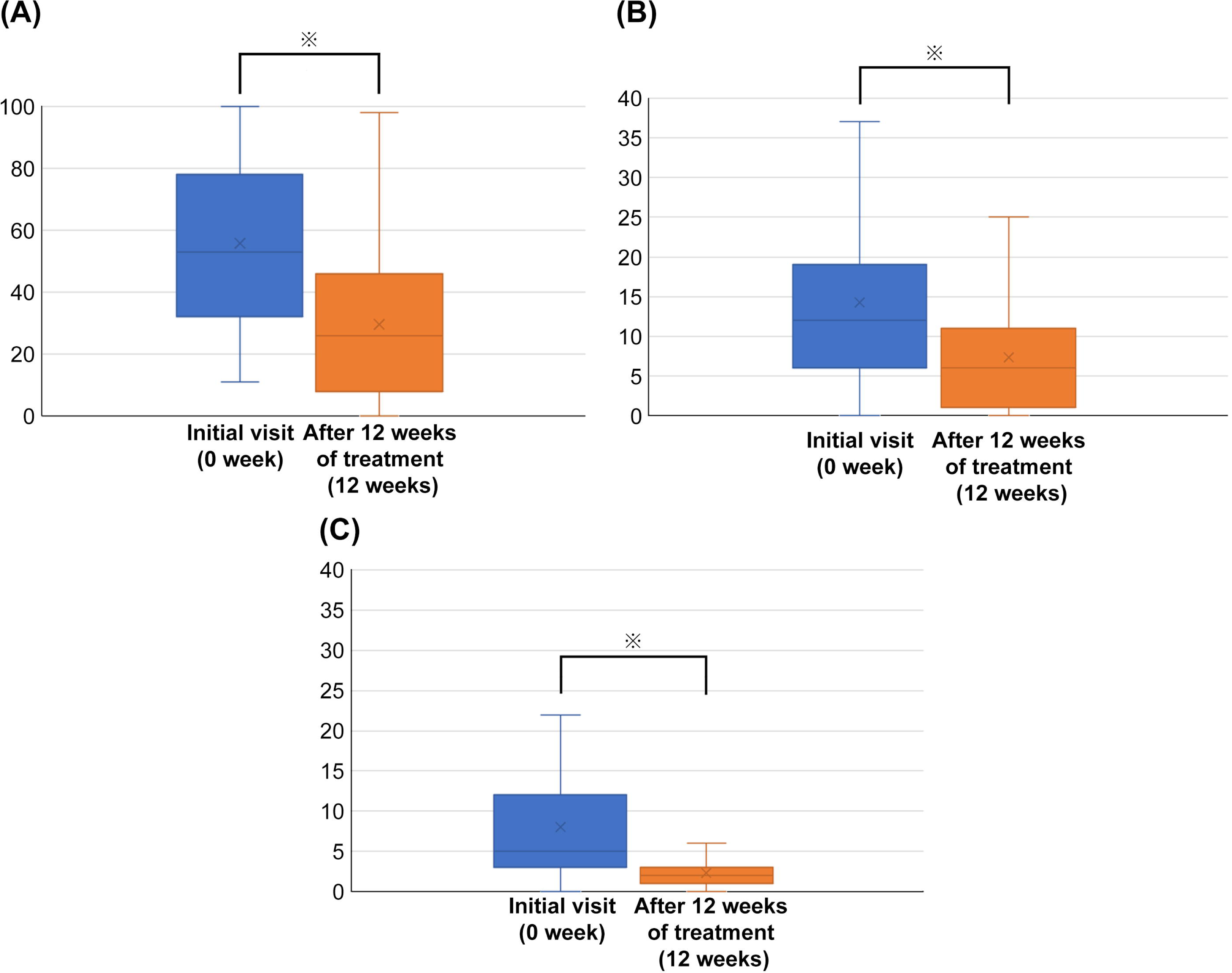
**Figure 2A. Comparison of VAS between at initial visit and after 12 weeks of treatment.** ※P<0.01 (0week vs 12weeks) **Figure 2B. Comparison of BDI between at initial visit and after 12 weeks of treatment.** ※P<0.01 (0week vs 12weeks) **Figure 2C. Comparison of HDRS between at initial visit and after 12 weeks of treatment.** ※P<0.01 (0week vs 12weeks)

## Discussion

This study showed that while PCS in patients with BMS or PIFP improved with treatment, it did not improve to NSV after 12 weeks of intervention, whereas MCS improved to the same level as NSV after 12 weeks of intervention. The results demonstrated that treatment intervention improved BDI and HDRS, a measure of depression. Emotional factors have been reported to be strongly involved in the deposition and relief of chronic pain and may have diverse effects on pain expression [20]. This study indicates that especially when no organic cause has been identified for physical symptoms, prompt collaboration with psychosomatic medicine and psychiatry is extremely important, rather than making a definitive diagnosis and treatment by dentistry alone.

The validity of the study results is considered high because the SF-8, a standard and universally used instrument worldwide, was used to measure QOL. Previous reports using the SF8 have included evaluation for rheumatism [21] and stroke [22], and the study results indicate that it can be used adequately in the field of dentistry. Visual assessment methods such as the VAS [23] and the face scale [24] have been previously used as assessment methods that include patients’ mental satisfaction; however, it is difficult to say that they are sufficient. To evaluate medical interventions, subjective factors including psychosocial aspects should be measured, and a simple scale is needed for this purpose. In other words, in addition to the subjective evaluation of physicians and dentists, improvement of patients’ ADL, and tracking the blood data, a scale similar to the SF8, which easily measures PCS and MCS, can be a good means of communication between physicians and patients.

Although the above findings are similar, MCS recovered while PCS did not recover to NSV in this study. Thus, emotional turbulence may cause muscle hyperactivity induced by the central nervous system, resulting in parafunctional habits [25]. Depression is also reported to be more likely a consequence than a precursor of living with pain so the mind needs to gain supremacy over the body to compensate for pain. In other words, the emotional side of pain should be managed first, rather than both the mental and physical sides, and our study may have been the result of intervention from the mental side. However, restoring PCS is still necessary to improve the patients’ QOL. This may be because many patients do not yet achieve the level where treatment can be terminated although symptoms tend to decrease after up to 12 weeks of treatment, and treatment for PCS may take a few more weeks to recover to a level comparable to NSV. One report revealed that it took 2 years for PCS to finally approach the national norm in a patient with postoperative head and neck cancer [26]. This finding suggests that PCS may not be adequately improved even with long-term follow-up.

MCS improves first due to “the effects of specialized psychiatric treatment with psychotherapy and antidepressants.”

This study has several limitations. First, the number of patients and the study duration are limited (as noted above, PCS may improve with longer follow-up), and it is a single-center study. In addition, as the SF-8 is a scale that can be used for a wide range of patients, from normal participants to patients with chronic diseases, it is not a disease-specific scale for chronic pain in the orofacial region.

In conclusion, we used the SF-8 Japanese version to investigate (1) the QOL of patients with BMS or PIFP (compared to a Japanese control group) and (2) whether the therapeutic intervention improves the QOL and pain reduction (comparison between 0 and 12 weeks). The results showed that therapeutic intervention improves MCS and reduces pain; however, improving PCS takes time.

## Data Availability

The data underlying the results presented in the study are available from Tatsuya Tokura.

## Acknowledgments

The authors would like to thank Enago (www.enago.jp) for the English language review.

